# Whole Genome Sequencing Analysis of Spike D614G Mutation Reveals Unique SARS-CoV-2 Lineages of B.1.524 and AU.2 in Malaysia

**DOI:** 10.1101/2021.08.11.21261902

**Authors:** Ummu Afeera Zainulabid, Aini Syahida Mat Yassim, Sharmeen Nellisa Soffian, Mohamad Shafiq Mohd Ibrahim, Norhidayah Kamarudin, Mohd Nazli Kamarulzaman, How Soon Hin, Hajar Fauzan Ahmad

**Affiliations:** Faculty of Industrial Science Technology, Universiti Malaysia Pahang, 26300 Gambang, Pahang, Malaysia; Department of Internal Medicine, Kulliyyah of Medicine, International Islamic University of Malaysia, 25200 Kuantan, Pahang, Malaysia; Department of Paediatric and Dental Public Health, Kulliyyah of Dentistry, International Islamic University Malaysia, 25200 Kuantan, Pahang, Malaysia; Department of Pathology and Laboratory Medicine, Kulliyyah of Medicine, International Islamic University of Malaysia, 25200 Kuantan, Pahang, Malaysia; Department of Surgery, Kulliyyah of Medicine, International Islamic University of Malaysia, 25200 Kuantan, Pahang, Malaysia; Centre for Research in Advanced Tropical Bioscience (Biotropic Centre), Universiti Malaysia Pahang, 26300 Gambang, Pahang, Malaysia

**Author notes:** Corresponding author (HFA) (HSH). These authors contributed equally to this work (Ummu Afeera Zainulabid, Aini Syahida Mat Yassim).

**Keywords:** SARS-CoV-2, Mutation, D614G, Spike Protein, G1223C, Clade, Malaysia, Pahang

## Abstract

The SARS-CoV-2 has spread throughout the world since its discovery in China, and Malaysia is no exception. WGS has been a crucial approach in studying the evolution and genetic diversity of SARS-CoV-2 in the ongoing pandemic, and while an exceptional number of SARS-CoV-2 complete genomes have since been submitted to GISAID and NCBI, there is a scarcity of data from Malaysia. This study aims to report new Malaysian lineages responsible for the sustained spikes in COVID-19 cases during the third wave of the pandemic. Patients whose nasopharyngeal and oropharyngeal swabs were confirmed positive by real-time RT-PCR with Ct-value < 25 were chosen for WGS. The 10 SARS-CoV-2 isolates obtained were then sequenced, characterized and analyzed, including 1356 sequences of the dominant lineages of D614G variant currently circulating throughout Malaysia. The prevalence of clade GH and G formed strong ground of the discovery of two Malaysian lineages that caused sustained spikes of cases locally. Statistical analysis on the association of gender and age group with Malaysian lineages revealed a significant association (*p* < 0.05). Phylogenetic analysis revealed dispersion of 41 lineages, for which 22 lineages are still active. Mutational analysis observed unique G1223C missense mutation in Transmembrane Domain of Spike protein. Thus, calls for the large-scale WGS analysis of strains found around the world for greater understanding of viral evolution and genetic diversity especially in addressing the question of the effect of deleterious substitution mutation in transmembrane region of Spike protein.

## Introduction

The emergence of severe acute respiratory syndrome coronavirus 2 (SARS-CoV-2) also known as COVID-19 in Wuhan, China in December 2019 resulted in an unprecedented global outbreak and has now become a major public health issue [1–4]. To date, more than 190,671,330 confirmed cases of COVID-19, including 4,098,758 deaths have been reported to the World Health Organization (WHO) worldwide [5]. By the same date, the cumulative number of confirmed cases of COVID-19 in Malaysia had reached 939,899, of which 7,241 died from the disease while 798,955 survived. The daily number of confirmed cases of COVID-19 has continued to soar with more than 10,000 cases per day since July 13, 2021 [6]. Malaysia is facing a much tougher task in curbing the COVID-19 pandemic in its third wave due to the emergence of outbreaks in Sabah [7]. Since then, the highest lineage contributor during the third wave of pandemic as B.1.524, with Glycine at residue 614 (G614) of the Spike protein, replacing Aspartic acid (D614) and Valine at residue 701 (V701) of the Spike protein, replacing Alanine (A701) [8].

The WHO defined SARS-CoV-2 Variant of Concerns (VOCs) as variants with clear evidence indicating significant impact in higher transmissibility, severity (including hospitalizations or death) and/or immunity due to significant reduction in neutralization by antibodies generated during previous infection or vaccination, as well as reduced effectiveness of treatments or vaccines that is likely to have an impact on the epidemiological situation [9, 10]. Whereas Variant of Interests (VOIs) are variants with specific genetic markers that have been associated with changes to receptor binding, reduced neutralization by antibodies generated against previous infection or vaccination, reduced efficacy of treatments, potential diagnostic impact, or predicted increase in transmissibility or disease severity [11]. The SARS-CoV-2 VOCs UK B.1.1.7 (Alpha) was first detected in Malaysia since February followed by VOIs Nigerian B.1.525 (Eta) on March, and VOCs South African B.1.351 (Beta) on March this year. VOCs Indian B.1.617.2 (Delta) were detected from June in Peninsular Malaysia, followed by VOIs B.1.617.1 (Kappa) from June this year [12]. In Sarawak, the first Delta variant was detected on June, together with VOIs Philippines P.3 (Theta) in 2021[13].

Of interest, all of the VOCs and VOIs detected in Malaysia share a D614G mutation in their Spike protein [14, 15]. Until recently, 90.30% of all COVID-19 infection in Malaysia has been due to the D614G variant, and this mutation remains in all new emerging variants [14]. As a result of positive natural selection, it was found that D614G increases the infectivity, viral fitness, transmission rate and efficiency of cellular entry for the SARS-CoV-2 virus across a broad range of human cell types [8,16–22]. Nevertheless, D614G mutation alone has not been shown to cause higher COVID-19 mortality or clinical severity, or alter the efficiency of the current laboratory diagnostic, therapeutics, vaccines or public health prevention strategies [10, 23]. Therefore, in this study, we analyzed the dominant lineages of D614G variants currently circulating in Malaysia using whole complete genome of the Malaysian SARS-CoV-2 deposited to the Global Initiative on Sharing All Influenza Data (GISAID) database. This study aims to report new Malaysian lineages that are responsible in causing sustained spikes in COVID-19 cases throughout the third wave of the pandemic in Malaysia. We also investigated the divergency of the D614G variant of the Pahang SARS-CoV-2 virus isolates and its possible origin. Here we report the list of amino acid mutations in Spike protein detected in Pahang SARS-CoV-2 strains and hence predict the changes in protein-protein binding affinity due to these missense amino acid mutations. Finally, we computationally predict a possible effect on the biological function of a Spike protein due to occurrence of a unique mutation of G1223C in Transmembrane (TM) Domain of Spike protein which only detected in Pahang SARS-CoV-2.

## Materials and Methods

### Sample selection

Nasopharyngeal and oropharyngeal swab test results from 10 patients that were confirmed positive for SARS-CoV-2 through real-time reverse transcriptase-PCR (real-time RT-PCR) at the local hospital was selected for whole genome sequencing.

### RNAs extraction

Samples were selected based on real-time RT PCR quality data where the RNAs must be more than 10ng/ul and have low cycle threshold. The RNA was extracted before real-time reverse transcriptase (RT-PCR) techniques to identify SARS-CoV-2. The genome information was then uploaded to international databases such as NCBI and GISAID.

### Next-generation sequencing of the full-length viral genome

A next-generation sequencing (NGS) library was constructed after amplifying the isolates’ full-length genes using synthesized cDNA from SuperScriptIV (ThermoFisher Scientific, USA) with some modifications [24, 25]. Briefly, 5 µl of the cDNA was used as template for multiplex PCR using Q5 polymerase (NEB, USA) as well as the Artic v3 primer pools during library preparation. The constructed library was sequenced on an iSeq 100 System (Illumina, USA) (with run configuration of 1 × 300 bp).

### Sequence analysis

The SARS-CoV-2 genome was reconstructed from the raw reads using a combination of bioinformatic tools as listed in https://github.com/CDCgov/SARS-CoV-2_Sequencing/tree/master/protocols/BFX-UT_ARTIC_Illumina. Genome sequences from other studies related to human and animal coronavirus were mined from the GISAID (https://www.gisaid.org) and NCBI GenBank (https://www.ncbi.nlm.nih.gov/genbank/).

### Public database SARS-CoV-2 genome analysis

For the study of dominant lineage and D614G frequency, a total of 1356 whole or complete genome sequences of Malaysian SARS-CoV-2 Malaysia that were submitted to GISAID were retrieved from March 1, 2020 to July 19, 2021. Analysis of lineage distribution and clade frequency were performed manually by using Pivot table in Excel. Real-time Malaysia SARS-CoV-2 Genomics Surveillance updates were monitored v*ia* (https://bit.ly/2UEFFGt).

The first virus from each lineage with D614G mutation in Spike protein was extracted using patient’s status metadata downloaded from GISAID until July this year. To do this, 1356 viruses were analysed manually using Pivot table and the date was filtered to months and year in Excel. The lineage description was classified according to the PANGO Lineage List (https://cov-lineages.org/lineage_list.html).

### Phylogenetic tree analysis

A total of 1005 complete whole genome sequences of Malaysian variant with D614G mutation were retrieved from GISAID database (S1 Table 1). A complete genome of Wuhan-Hu-1 (NC_045512) was downloaded from GenBank (https://www.ncbi.nlm.nih.gov/sars-cov-2/) for outgroup. The multiple sequence alignment was performed using DECIPHER [26] and SeqinR [27] packages in R version 4.0.2 and finalized using MEGA X 11 [28].

**Table 1.** The distribution of the lineage between gender, patient status and age groups.

| | | Lineage, n (%) | | Total, n (%) | $\chi^2$ | p value |
| --- | --- | --- | --- | --- | --- | --- |
|  |  | AU.2 (GH) | B.1.524 (G) |  |  |  |
| Gender | Male | 134(37.9) | 220(62.1) | 354(100) | 3.862 | 0.049 |
|  | Female | 132(45.5) | 158(54.5) | 290(100) |  |  |
| Patient Status | Deceased | 0 | 3(100) | 3(100) | 1.541 | 0.463 |
|  | Hospitalized | 0 | 6(100) | 6(100) |  |  |
|  | Live | 36(14.7) | 209(85.3) | 245(100) |  |  |
| Age Groups | 0-14 years | 27(50.9) | 26(49.1) | 53(100) | 6.542 | 0.038 |
|  | 15-64 years | 190(40.7) | 277(59.3) | 467(100) |  |  |
|  | >64 years | 43(54.4) | 36(45.6) | 79(100) |  |  |

Evolutionary analyses conducted in MEGA X was inferred using the Neighbor-Joining (NJ) method [29]. The bootstrap consensus tree inferred from 500 replicates was taken to represent the evolutionary history of the taxa analyzed. Branches corresponding to partitions reproduced in less than 50% bootstrap replicates were collapsed. The evolutionary distances were computed using the Kimura 2-parameter method and are in the units of the number of base substitutions per site. The rate variation among sites was modelled with a gamma distribution (shape parameter = 1). All ambiguous positions were removed for each sequence pair (pairwise deletion option).

### Mutation analysis *via* computational prediction tools

Mutation analysis were analyzed using Nextclade v.1.5.2, a web-based analysis server (https://clades.nextstrain.org) by comparing against a wild-type of Wuhan-Hu-1 (NC_045512.2).

To evaluate the effect of mutations on protein-protein binding affinity, a 3D structure model of wild-type Spike protein (<u>YP_009724390.1</u>) was first generated using SWISS-model based on the most fitted protein template PDB ID: <u>6XR8</u> (Distinct conformational states of SARS-CoV-2 spike protein. This 3D structure model however covers a residue of 14-1162 only. For analyzing the effect of amino acid substitution in TM domain, a 3D structure PDB ID: <u>7LC8</u> (SARS-CoV-2 Spike protein TM domain) was used. Both 3D structure model of YP_009724390.1 and 7LC8 were uploaded to mCSM-PPI2 server [30]. Next, the potential pathogenicity effect of the amino acid substitution on TM domain biological function was investigated by uploading a 3D structure of TM domain PDB ID: 7LC8 and TM domain amino acid sequence onto mCSM-membrane [31] and uploading TM amino acid sequence onto Protein Variation Effect Analyzer (PROVEAN) [32] and SNAP 2 tools [33]; the web-based servers for predicting the effect of mutations on the biological function of a protein. The servers predicted the consequence of amino acid mutation to be whether benign or pathogenic, deleterious or neutral, effect or neutral, respectively.

### Statistical analysis

Data are presented as count and percentage. Chi-square test were carried out using IBM SPSS v25.0 to testing the statistical significance association of gender, patient status and age groups with Malaysian lineages. All level of significances were set at *p* < 0.05.

## Results

### The evolution of D614G variant of SARS-CoV-2 in the Malaysian population

Of the1,502 SARS-CoV-19 complete genomes deposited to GISAID database, 1,356 contained Spike D614G mutation in their genomes. To better characterize the local distribution of lineages that may contribute to the constant increase in COVID-19 cases in Malaysia, Fig. 1 summarizes the distribution of the D614G variant lineages throughout the country since it was first detected. Based on the GISAID database analysis, there were 41 lineages of D614G variant dispersed throughout Malaysia. Lineage B.1.524 (n= 419) and AU. 2 (n=311) appears to have caused significant transmission of the virus locally compared to Variant of Concern (VOC) Alpha B.1.1.7 (n=11), Beta B.1.351(n=161), B.1.351.3 (n=2), and Delta B.1.617.2 (n=58); Variant of Interest (VOI) Eta B.1.525(n=3), Kappa B.1.617.1(n=3), as well as lineages currently designated alerts for further monitoring, such as P.2 (n=1), P.3 (n=10), B.1.466.2 (n=70), B.1.214.2 (n=1).

**Fig. 1.**
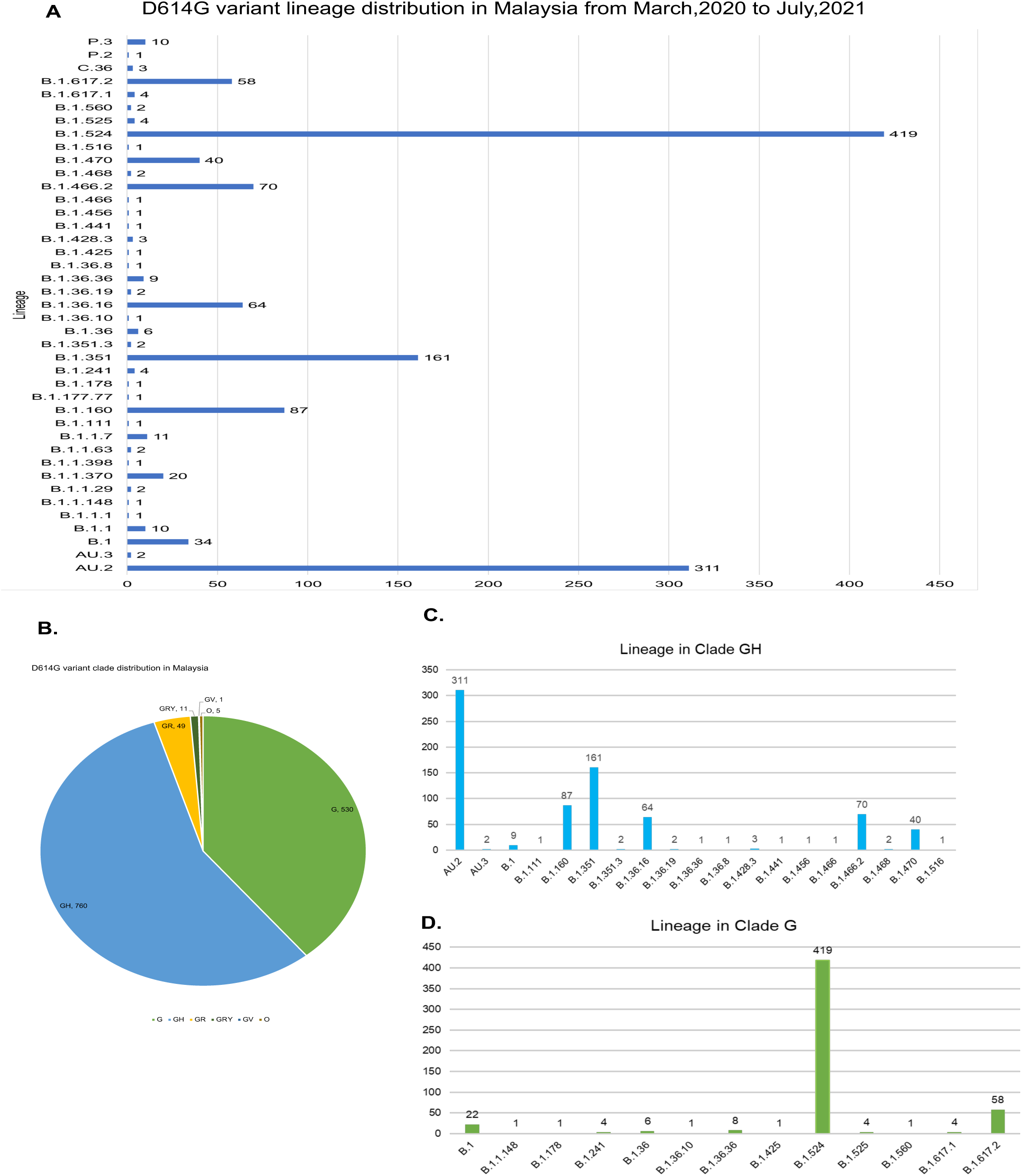
D614G variant lineage and clade distributions in Malaysia. A. Distribution of lineages of D614G variant based on all complete genomes from Malaysia deposited in GISAID until July 5, 2021 (n=1356). B. D614G variant clade distribution based on all complete genomes from Malaysia deposited in GISAID until July 19, 2021. (n=1356) C. Lineage clustered to clade GH. D. Lineage clustered to clade G.

Next, we investigated the frequency of D614G variant clades circulating in Malaysia until July this year. It appeared that of the six D614G variant clades (GH, G, GR, GRY, O and GV), GH makes up the largest clade with 760 of genomes from different lineages (Fig. 1B). Further analysis of Clade GH shows lineage AU.2 had appeared most often in the transmission of the disease (Fig. 1C) followed by clade G, in which lineage B.1.524 seems to be the highest contributor in the local transmission of Covid19 (Fig. 1D).

Our findings on genomic surveillance in depicting local transmission and evolution of the D61G variant revealed that two of the variants had emerged locally: B.1.524, and AU.2. Of these two, B.1.524 had silently caused the largest local transmission of the D614G variant in Malaysia (n= 419), followed by AU.2 (n=311).

Furthermore, splitting the genomes analysis based on years, we found a clear pattern of lineage distribution which demonstrates how the major lineages disperse throughout Malaysia in 2020 and 2021 (S1 Fig 1). While the B.1.524 may have contributed heavily to the initial number of D614G lineage actively spreading locally, data suggest the AU.2 lineage, is currently taking its place as the major D614G variant contributor in spreading the disease. This raised a question of where these Malaysia lineages originated from. Based on our current analysis, we suggest that AU.2 might have originated from Sarawak, while B.1.524 remains unknown.

Next we analysed the association of gender, patient status and age group with Malaysian lineages B.1.524 and AU.2. According to Table 1, there is significant association in lineage between the gender and age group. The association lineage between males and females shows significant association (62.10% vs 54.50%, *p* = 0.049). It is also observed there is significant association between lineage and age groups (49.10% vs 59.30% vs 45.60%, *p* = 0.038). However, there is no significant association between lineage and patient’s status in term of disease severity.

### Origin of the massive spread of COVID-19 cases in Pahang

To infer the origin of the D614G variant that was responsible in causing widespread COVID-19 infections in Pahang this year, we built a NJ phylogenetic tree using D614G variant complete genomes downloaded from GISAID (Fig. 2). The virus sample collection dates restricted to January 1, 2021 until July this year (n=1005). Based on our analysis, there were 22 SARS-CoV-2 lineages actively dispersed in Malaysia this year. NJ phylogenetic analysis constructed in Fig. 2A demonstrated that despite being identified as Malaysian lineages, both AU. 2 and B.1.524 were distantly diverged from each other.

**Fig. 2.**
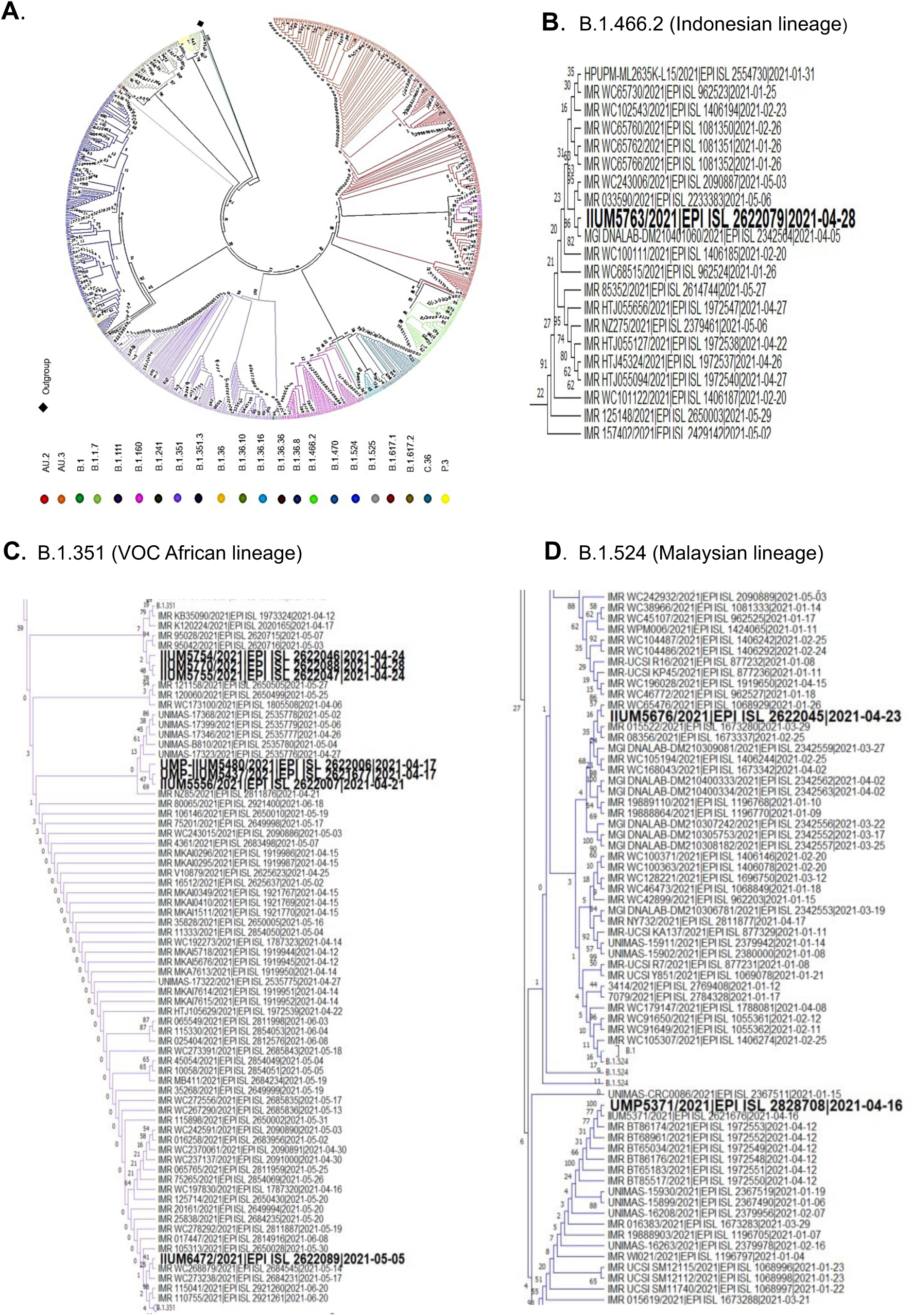
Phylogenetic tree of 1005 complete genomes of Malaysia D614G variant in 2021. A. The evolutionary history was inferred using the Neighbor Joining (NJ) method with 500 bootstrap replications. Branches corresponding to partitions reproduced in less than 50% bootstrap replicates are collapsed. The percentage of replicate trees in which the associated taxa clustered together in the bootstrap test (500 replicates) are shown next to the branches. The evolutionary distances were computed using the Kimura 2-parameter method and the rate variation among sites was modeled with a gamma distribution (shape parameter = 1). This analysis involved 1005 nucleotide sequences. Codon positions included were 1st+2nd+3rd+Noncoding. All ambiguous positions were removed for each sequence pair (pairwise deletion option). There was a total of 29672 positions in the final dataset. Evolutionary analyses were conducted in MEGA X. B. A closed-up view of NJ phylogenetic tree focusing on Pahang D614G variant of IIUM 5763/2021, from lineage B.1.466.2. C. A closed-up view of NJ phylogenetic tree focusing on Pahang D614G variant of IIUM5754/2021, IIUM5770/2021, IIUM5755/2021, IIUM-UMP5480/2021, IIUM-UMP5437/2021, IIUM5556/2021 and IIUM6472/2021, from lineage B.1.351. D. A closed-up view of NJ Phylogenetic tree focusing on Pahang D614G variant of UMP5371/2021 and IIUM5676/2021, from lineage B.1.524.

In order to generate comprehensive phylogenetic analyses of the D614G variant actively spreading in Pahang, a closed up view of the evolutionary tree constructed in Fig. 2A was visualized in Fig. 2B-2D. A closed up view of the NJ phylogenetic tree shown in Fig. 2B is focused on IIUM5763/2021, clustered to B.1.462 (Indonesian lineage). While Fig. 2C highlighted a phylogenetic tree analysis of IIUM6472/2021, UMP-IIUM5437, UMP-IIUM5480, IIUM5556/2021, IIUM5754/2021, IIUM5755/2021, IIUM5770/2021, clustered to Beta B.1.351. And Fig. 2D visualized the result of evolutionary analysis of IIUM5371/2021and IIUM5676, clustered to B.1.524 (Malaysian lineage). Further analysis of Fig. 2B suggested that IIUM 5763/2021 was closely related to sample MGI DNALAB-DM210401060/2021, which originated in Selangor, including IMR WC243006/2021 and IMR 033590.

IIUM 5770/2021. IIUM 5755/2021 and IIUM 5754/2021 shown in Fig. 2C were strongly related to one another as the viruses were sampled from patients who had travelling history from similar place. Remarkably, these three genomes demonstrated distant correlations with other virus genomes deposited in the GISAID database. Comparison of genomes IIUM 5556/2021 and UMP-IIUM 5437/2021 show highest similarity to each other, and are closely related to UMP-IIUM 5480/2021. Nevertheless, these three genomes were distantly diverged to IIUM 6472/2021 although they share the same initial internal node on the NJ tree. Our analysis demonstrated that IIUM 6472/2021 was closely related to IMR_WC268879/2021, which was sampled locally.

Referring to Fig. 2D, UMP 5371/2021 was distantly diverged to IIUM 5676/2021 with different internal nodes. It appears that IIUM 5676/2021 was closely related to IMR WC65476/2021 sampled from an unspecified location in Malaysia. UMP5371/2021 was found to be closely related to IMR BT86174.

### Mutations in Spike protein of Malaysian lineages

419 complete genomes of B.1.524 and 311 complete genomes of AU.2 were uploaded to Nextclade v.1.5.2 (https://clades.nextstrain.org) to analyse dominant mutations occur in Spike protein of Malaysian lineages. Based on our analysis presented in Fig. 3, other than D614G mutation, B.1.524 carries A701V mutation in Spike protein. Whereas, AU.2 carries a mutation at positions 439 from Asparagine (N) to Lysine (K), 681 from Proline (P) to Arginine(R) and 1251 from Glycine (G) to Valine (V).

**Fig. 3:**
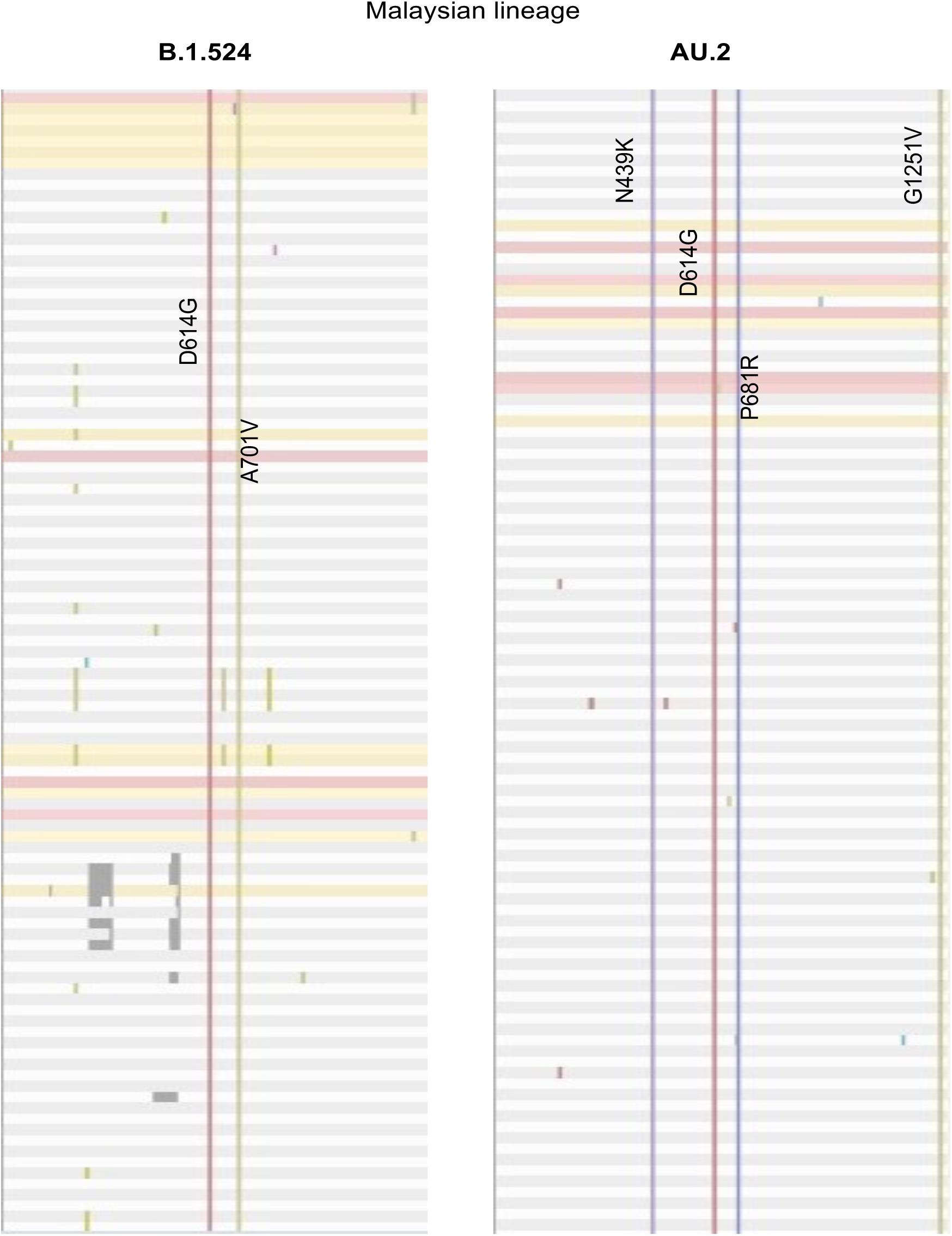
List of mutations in Spike protein of Malaysian lineages. A. B.1.524 carries a mutation of D614G and A701V. B. AU.2 carries a mutation of N439K, D614G, P681R and G1251V. A screen shot images presented here represent some of genomes analysed using Nexclade v.1.5.2 (https://clades.nextstrain.org).

### Amino acid mutations in Spike protein of the Pahang-D614G variant SARS-CoV-2

1005 complete genomes of the D614G variant were analysed for sequence quality and mutation. Of that, 986 complete genomes passed Nextclade’s sequence quality control. Focusing on mutation in the Spike protein, our analysis revealed that all of Pahang’s SARS-CoV-2 isolates had a unique substitution mutation of Glycine (G) to Cysteine (C) at position 1223 (G1223C) which was not found in the other 976 genomes (S4 Fig. 2). A list of amino acid mutations in Spike protein of Pahang’s G614 variant were summarized in Fig. 4.

**Fig. 4:**
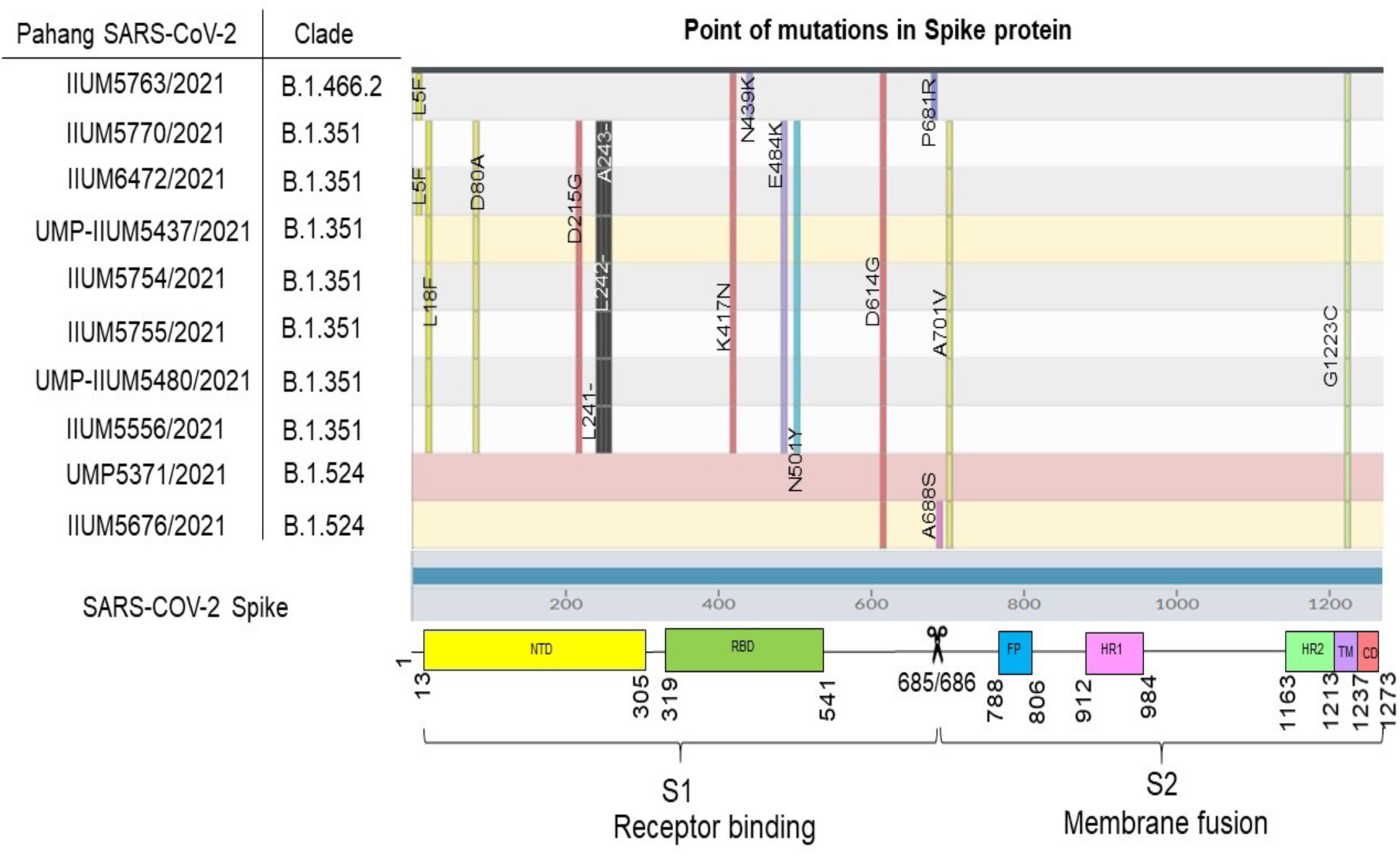
Point of mutations in Spike protein of Pahang SARS-CoV-2, D614G variant with a schematic representation of the SARS-CoV-2 Spike shown at the bottom. The listed domain boundaries was defined according to recently published source. NTD: N-terminal domain; RBD: receptor-binding domain; 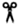 :S1/S2 cleavage site; FP: fusion peptide; HR1:heptad repeat 1; HR2: heptad repeat 2; TM: transmembrane domain; CD: connector domain. An amino acid mutation was analysed using Nexclade v.1.5.2 (https://clades.nextstrain.org).

The effects of single-point mutations on protein-protein interaction binding affinity was performed using mCSM-PPI2. The result of the analysis was summarized in Table 2. To do this, a 3D structural model of wild type Spike protein (YP_0097243901) was first generated through SWISS MODEL using a protein template model of 6XR8 (distinct conformation states of SARS-CoV-2 Spike protein). The 3D structure model of YP_0097243901 generated only covered residues 14 to 1162. As such, prediction of the effects of missence mutation on protein-protein interaction binding affinity only covered residues within this region. Of note, mCSM-PPI2 is unable to predict the change in protein interaction affinity in single amino acid deletions, hence analysis on L241del, L242del and A243del were not included in Table 2. To analyse the effects of G1223C mutation in the TM region of Spike protein, a 3D structure model of the SARS-CoV-2 Spike protein TM domain 7LC8 downloaded from RCSB Protein Data Bank was directly uploaded to the mCSM-PPI2 server. The effects of G1223C mutation in the TM domain was included in Table 2. Together, the missence mutations L18F, N501Y, A701V and G1223C seems to have increased the binding affinity of the Spike protein, while mutations D80A, D215G, K417N, N439K, E484K and A688S had the opposite effect. Focusing on the unique mutation G1223C, the difference in intramolecular interactions between the wild type G1223 and the mutant C1223 shown in Fig. 3, suggests that G1223C does not cause significant structural rearrangement of the TM domain, except for the gain in salt bridge between C1223 and G1219 (Fig. 5 - Mutant (C1223)).

**Fig 5.**
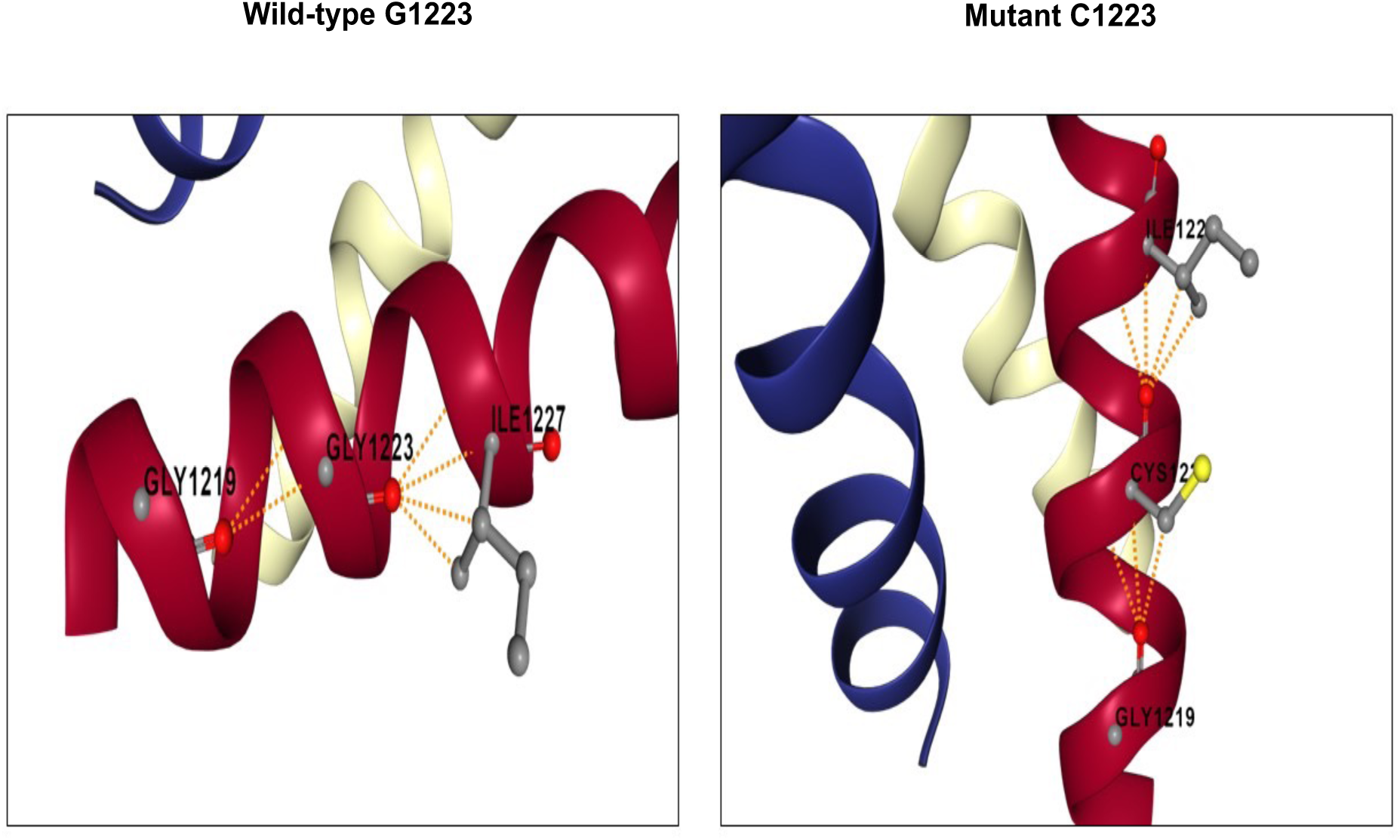
Interatomic interaction in TM domain of wild-type (G1223) compared to mutant (C1223). The surrounding residues with close interaction with the wild-type and mutant residue of 1223 are highlighted.

**Table 2.**
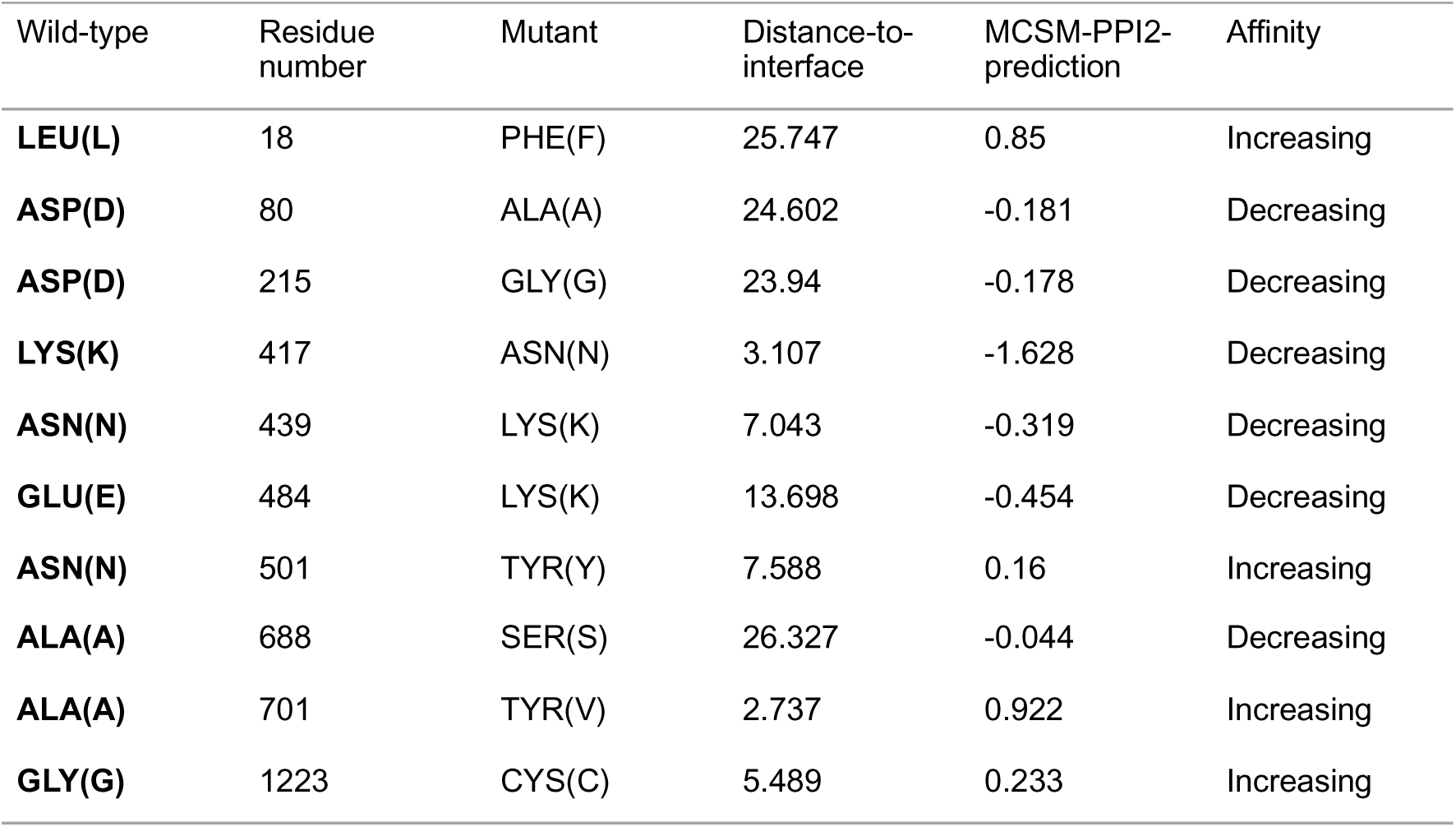
The effect of missenses mutation in Spike protein of Pahang D614G SAR-CoV-2 as predicted by mCSM-PPI2.

| Wild-type | Residue number | Mutant | Distance-to-interface | MCSM-PPI2-prediction | Affinity |
| --- | --- | --- | --- | --- | --- |
| **LEU(L)** | 18 | PHE(F) | 25.747 | 0.85 | Increasing |
| **ASP(D)** | 80 | ALA(A) | 24.602 | -0.181 | Decreasing |
| **ASP(D)** | 215 | GLY(G) | 23.94 | -0.178 | Decreasing |
| **LYS(K)** | 417 | ASN(N) | 3.107 | -1.628 | Decreasing |
| **ASN(N)** | 439 | LYS(K) | 7.043 | -0.319 | Decreasing |
| **GLU(E)** | 484 | LYS(K) | 13.698 | -0.454 | Decreasing |
| **ASN(N)** | 501 | TYR(Y) | 7.588 | 0.16 | Increasing |
| **ALA(A)** | 688 | SER(S) | 26.327 | -0.044 | Decreasing |
| **ALA(A)** | 701 | TYR(V) | 2.737 | 0.922 | Increasing |
| **GLY(G)** | 1223 | CYS(C) | 5.489 | 0.233 | Increasing |

Subsequently, further analysis on the effects of all missense mutations occurring at the 1223 residue on the TM domain functionality was performed using SNAP2, PROVEAN and mCSM-membrane was summarized in Table 3. Overall, two computational prediction tools, PROVEAN and SNAP2 suggesting for a possibly serious evolutionary damaging consequence of protein biological function due to G1223C mutation in TM domain.

**Table 3.** Prediction of change in TM biological function.

| **Mutation in TM** | **mCSM-membrane**<br>**(Benign/Pathogenic)** | **PROVEAN**<br>**(Neutral/Deleterious)** | **SNAP 2**<br>**(Neutral/Effect!)** |
| --- | --- | --- | --- |
| **G1223C** | Benign | Deleterious | Effect |

Legends: Computational prediction was performed by using three web-based tools:
mCSM-membrane http://biosig.unimelb.edu.au/mcsm_membrane/
PROVEAN http://provean.jcvi.org/seq_submit.php
SNAP 2 https://roslab.org/services/snap2web/

## Discussion

The first incidence of COVID-19 in Malaysia was discovered on January of 2020 and was traced back to be originated from China [34]. The local Malaysian authority quickly developed standard guidelines for the management of COVID-19, including the set-up of designated hospitals and screening centers in each state [34]. To date, more than 951,884 COVID-19 cases was recorded, with increasing fatalities. Based on earlier report, we found that D614G mutation sample had been circulating in Pahang [24] and subsequently found elsewhere throughout Malaysia as the infection continues. For the record, the earliest study on SARS-CoV-2 virus genomes in Malaysia did not found D614G mutation, even the lineage B.6 that caused major outbreak for the second wave in Malaysia indicated with no D614G mutation in Spike protein [35].

Although major concerns have been raised on the emergence of Beta B.1.351 (VOC African lineage) and Delta B.1.617.2 (VOC Indian lineage) for their capability to cause severe infection and fatality, our detailed analysis of SARS-CoV-2 genomes from the Malaysian population reported two lineages of D614G variant that are actively dispersed locally. To our knowledge, the emergence of lineage B.1.524 was first detected on September last year. The analysis from early 2021 suggests that AU.2 had become dominant lineage that actively spread in Malaysia for 2021 followed by B.1.524. We observed that AU.2 had closest distance branch to B.1.4662 (Indonesian lineage), as both lineages carry the same amino acid mutations at N439K in RBD region and P681R in non RBD region of Spike protein. Its presence in Malaysia could be due to transference of the disease *via* visitors from Indonesia. Our detail analysis also suggests, AU.2 is not correlated to B.1.524. as B.1.524 harbours different mutations in Spike protein; A701V. Moreover, lineages assignment using pangolin (v2.1.6, https://github.com/cov-lineages/pangolin) described AU.2 and B.1.524 as Malaysia 94.0%, Indonesia 5.0%, United States of America 0.0%, India 0.0%, Singapore 0.0%, and Malaysia 76.0%, Singapore 16.0%, Thailand 3.0%, Philippines 2.0%, India 1.0%, respectively.

Of the 41 lineages of D614G variants detected in Malaysia, 19 lineages have disappeared, leaving 22 lineages still actively spreading into 2021. When a virus mutates, it undergoes alterations that may result in the development of new isolates following replication. Non-synonymous substitutions are extremely important since they result in an amino acid change, which in turn induces structural change [36]. This will later cause functional instability in isolates, resulting in susceptible illnesses, and even an increase in the degree of pathogenicity [36]. In nations with poor containment capability, it was proven that the SARS-CoV-2 mutant lineage G (S-D614G) was able to replace earlier lineages more efficiently and was associated with a higher degree of disease severity [37]. Moreover, the emergence of more virulent strains such as VOC and VOI that harbored the D614G mutation in Spike protein suggests that D614G variant had constantly subjected to positive selection pressure. Consequently, combination of various mutations in Spike protein had increased viral transmission [38–40], increased severity based on hospitalization cases and fatality rates [41], reduced susceptibility to the monoclonal antibody treatment [42] and reduced neutralization by convalescent and post vaccination sera [43–47].

Even though recent study suggested that the GR was a predominant clade in Asia [48], our study found that GH is the major infecting clade in Malaysia, follows by G. To the best of our knowledge, study related to AU.2 lineage in relating to disease epidemiology and pathology is scarce, however, the VOC of African B.1.351 that grouped together with AU.2 clade, was reported to significantly correlated with high disease severity and mortality [48]. Majority of mutations observed in this are deleterious in nature, unstable with impaired biological functions [49]. Based on these reasons, we anticipated that this may cause of high-risk transmission in Malaysia. On the other hand, the Malaysian lineage of B.1.524 is assigned to clade G that commonly associated with mild symptoms or asymptomatic cases [48]. Moreover, other study reported that the infection with clade G was not related with disease severity, and there was no clear indication of enhanced transmissibility despite greater viral loads [50].

We further analyzed the SARS-COV-2 genomes based on stratification of gender, age groups and disease’s status that available from the GISAID. Our metadata analysis showed higher (*p* < 0.05) prevalence among male patient with B.1.524 (G clade) variants. We postulate that male population are more vulnerable and susceptible to B.1.524 variant and a large-scale data is needed to understand further regarding this matter. Previous study showed that men with COVID-19 are more at risk for worse outcomes and mortality regardless of age [51] due to fundamental differences in the immune response between males and females despite socio-economic factors [52]. The disease distribution is significantly higher among adolescent and adult age group in both AU.2 and B.1.524 group (*p* < 0.05). This could be explained due to presence of comorbidities, immunological senescence and changes in ACE2 receptors [48]. However, our study showed negative correlation between both lineages in relation to disease severity. Here, we anticipate that this may be due to lack data related to the disease severity among Malaysian patients in GISAID.

Higher infectivity of the SARS-CoV-2 is associated with increased in binding affinity between Spike protein and ACE2 due to K417N, E484K, N439K and N501Y mutations in the RBD of Spike protein. While N501Y mutation alone enhanced Spike RBD-ACE2 affinity[53], combination of K417N, E484K and N501Y mutations in B.1.351 African lineage resulted in the highest degree of conformational alterations of RBD when bound to ACE2 [54–56]. Although N439K mutation in RBD was first found in already extinct lineage B.1.1.41 (also known as Engligh lineage), a new lineage B.1.258 (also known as UK lineage) independently acquired N439K[57]. While it is unknown whether B.1.466.2 (also known as Indonesian lineage) acquired N439K independently, AU.2 of Malaysian lineage is resembling to this lineage. Of concern, N439K mutation promotes evasion of antibody-mediated immunity by conferring resistance against several neutralizing monoclonal antibodies and reduces the activity of some polyclonal sera from patients recovered from infection [58]. However, there is no evidence of change in disease severity in a large cohort of patients infected with N439K virus [58].

In addition, A701V mutation at adjacent to the furin cleavage site of Spike protein subunit S1 and S2 in B.1.524 of Malaysian lineage was also occurs in B.1.351, and several variants under monitoring such as B.1.351+P384L (African), B.1.351+E516Q (Unclear) and Iota B.1.526 (USA) [59]. Although our computational prediction effect of A701V mutation suggest increase in protein-protein binding affinity, there is none functional experiment as we concern had been run to determine possible effect of A701V on virus transmissibility, immunity or infection severity. While A701V is still of unknown significance, further study should be performed accordingly.

In tracking the distribution of the ten lineages which caused blooming of positive COVID-19 cases in Pahang this year, it appears that all virus collected from Pahang have the same substitution of amino acid at 1233 from Glycine to Cysteine that related to mutation in TM domain of Spike protein that not common in Malaysia. Of interest, G1223C mutation had been found in England (13), Michigan (8), Stockholm (2), Portugal (1) and Germany (1) [60]. While the significance of G1223C mutation is still unknown, it is well known that Spike protein mediates entry of SARS-CoV-2 into target cells through two steps. First, it involves binding of RBD to its receptor human ACE2 and is proteolytically activated by human proteases at the S1/S2 boundary. Second, it follows by S2 of which include TM domain will undergoes structural change to mediate viral membrane fusion with targeted cells [61, 62]. To date, very little attention was put on the TM domain in the requirements for SARS-CoV cell entry. Although sequence analysis on TM domain among all coronaviruses Spike protein conducted previously [61, 63] revealed a high conservation rate, extensive mutation in TM domain of SARS-CoV however caused incapability of the virus to establish complete membrane fusion process [61]. Highly conserved small amino acids in TM domain of SARS-CoV-2 Spike protein (G1219, A1222, G1223, A1226) which initially thought to be important for TM domain oligomerization, but latest finding [63] showed neither glycine nor alanine in the trimer structure appear to be important for hydrophobic core formation. Thus, suggesting a possible role of the glycine motif is in a later step of fusion. We believe the effect of G1223C mutation in TM domain deserve to further analyze in future functional experiments for addressing above question.

This study has some limitations. First, the work on WGS in characterizing the circulating variants in Malaysia needed to be underscored systematically by representing Malaysian cases. Therefore, in order to success in combating the spread of COVID-19 in Malaysia, utilizing a viral genomics sequencing is critical to be used as a key tool for understanding the spread of COVID-19. By integrating viral genomics with epidemiological and modelling data, local transmission chains and regional spread were able to be tracked and audited in real time [64]. This strategy was proven to curb the spread of COVID-19 in a developed country for example Australia [65] and New Zealand [64]. Second, lack of patient clinical status details deposited to GISAID database hampered the analysis of the impact of the distribution of each individual clades on the disease epidemiology locally. Therefore, specifying whether the virus samples were collected from asymptomatic or mild symptoms, to severe or deceased might help to identify the prevalence of each major clade and lineage frequently detected. We also discovered a plethora of unclear entries that offer very little information about the real source of a sample. All of these issues can affect the effectiveness and accuracy of association studies. We therefore advocate for SARS-CoV-2 genomic data providers to comprehensively when submitting metadata, and encourage genomic database maintainers to be aware of potential errors in incoming samples and to actively support metadata standards. One option may be to entirely disregard samples with suspected metadata issues, however this may result in considerable reduction of sample size, thereby reducing the power of statistical studies [66].

## Conclusion

Here, we reported the most prevalent SARS-CoV-2 lineages of B.1.524 and AU.2 that sustained major outbreak of COVID-19 transmission during third wave of infection in Malaysia. Whereby the mutation at G1223C is under reported and further functional experiment is warranted. Furthermore, the N439K mutation that observed in RBD of AU.2 is deserved for comprehensive attention and monitoring due to its capability to increase virus infectivity while evading antibody-mediated immunity. Uncontrolled, and intensive virus transmission will result in the development of additional virus mutations, which may have significant influence on vaccination efficacy and perhaps, disease severity. The continual emergence of novel SARS-COV-2 variations highlights the need for public compliance with SOPs and other recommendations, notably mask use, hand cleanliness and physical separation, as well as the necessity to acquire herd immunity through the vaccination program.

These measures will aid in slowing viral transmission and reducing the likelihood of new variations emerging in the population.

## Data Availability

The sequences from WGS effort of this study were deposited in GenBank under the
accession numbers MW079428.1 and MZ443817.1 to MZ443824.1. The accession
numbers in the NCBI Sequence Read Archive (SRA) are SRP286590 and
SRP324679. All the SARS-CoV-2 with D614G mutation of Malaysia were collated from
GISAID (https://www.gisaid.org/).

## Acknowledgement

We acknowledge the COVID-19 task forces from Sultan Ahmad Shah Medical Centre @ IIUM and Universiti Malaysia Pahang, Malaysia.

## Supporting information

**S1 Fig 1. D614G variant lineage distribution in 2020 and 2021 based on complete genomes deposited to GISAID (Malaysia)**. A. The distribution of lineages from March to December, 2020. B. The distribution of lineages from January to July, 2020

**S2 Fig 2. Point of mutations in Spike protein of Pahang SARS-CoV-2, D614G variant compared to other genomes in Malaysia.** The screen shot image presented here represent some of the genomes analyzed using Nexclade v.1.5.2 (https://clades.nextstrain.org).

