## Supplementary material for "Whole Genome Sequencing Analysis of Spike D614G Mutation Reveals Unique SARS-CoV-2 Lineages of B.1.524 and AU.2 in Malaysia": S1 Fig 1

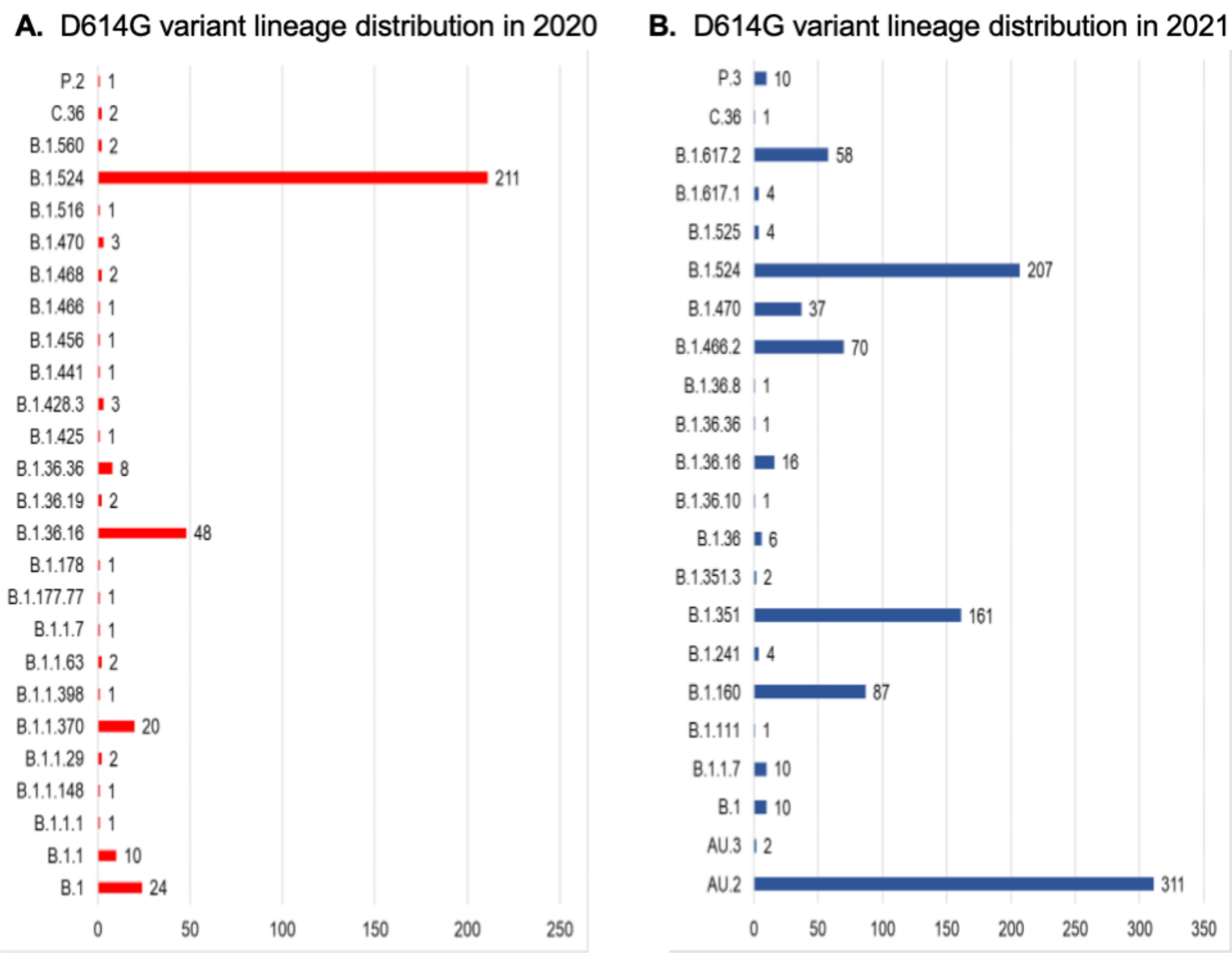


**S3 Fig 1. D614G variant lineage distribution in 2020 and 2021 based on complete genomes deposited to GISAID (Malaysia). A**. The distribution of lineages from March to December, 2020. **B.** The distribution of lineages from January to July, 2020
