## Supplementary material for "Whole Genome Sequencing Analysis of Spike D614G Mutation Reveals Unique SARS-CoV-2 Lineages of B.1.524 and AU.2 in Malaysia": S2 Fig 2

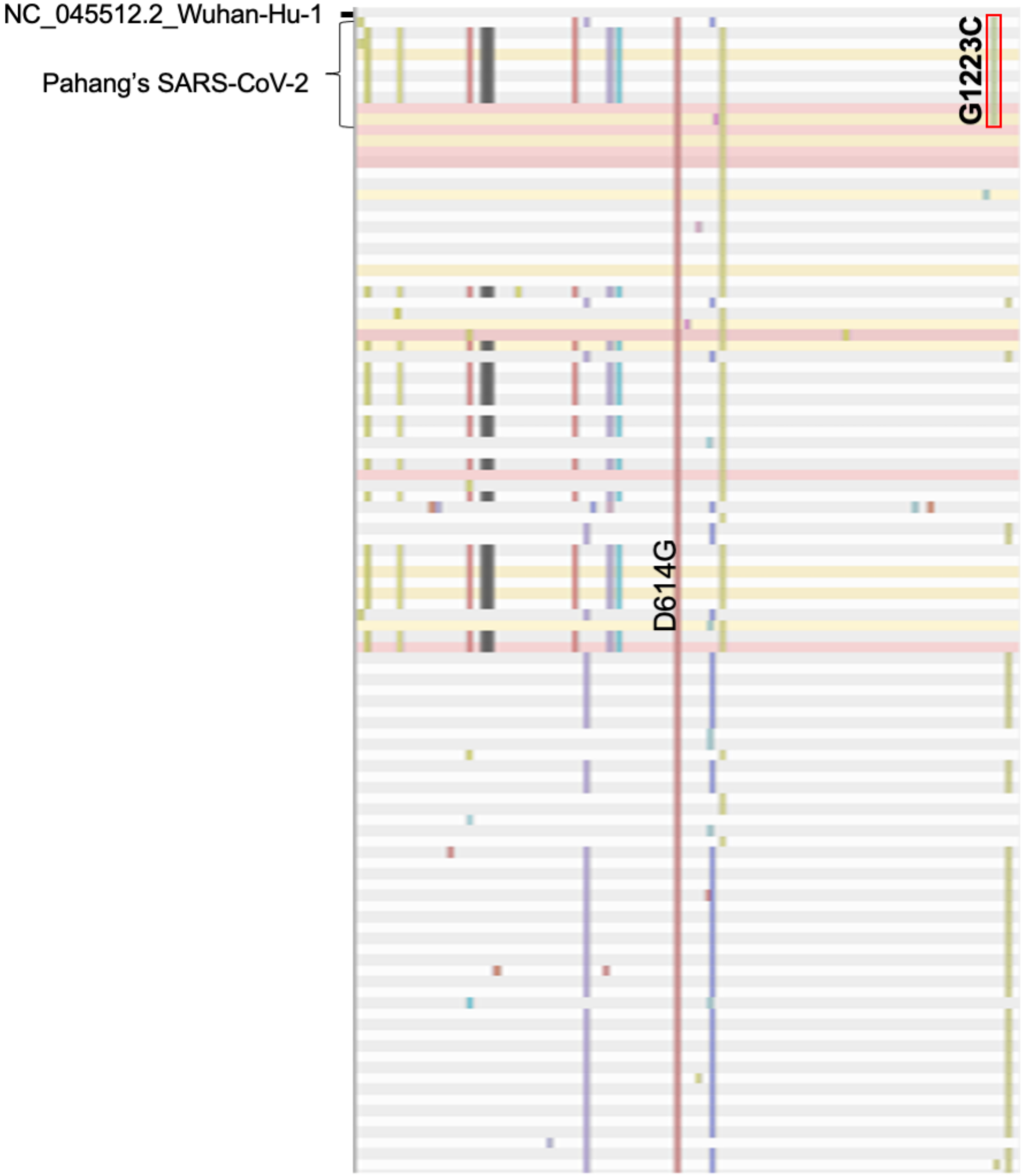


**S4 Fig. 2: Point of mutations in Spike protein of Pahang SARS-CoV-2,D614G variant compared to other genomes in Malaysia. The screen shot image presented here represent some of the genomes** analysed using Nexclade v.1.5.2 (https://clades.nextstrain.org).
